# Clinical significance of PD-1-binding soluble PD-L1 and MMPs in the microenvironment of gastric cancer and non-small cell lung cancer treated with PD-1/PD-L1 blockade

**DOI:** 10.1101/2024.01.06.23300672

**Authors:** Fumihiko Ando, Takeru Kashiwada, Shoko Kuroda, Takenori Fujii, Ryotaro Takano, Yoshishige Miyabe, Takashi Sakatani, Tomoko Asatsuma-Okumura, Masaaki Hashiguchi, Yoshikazu Kanazawa, Ryuji Ohashi, Hiroshi Yoshida, Masahiro Seike, Akihiko Gemma, Yoshiko Iwai

## Abstract

**Purpose:** The tumor microenvironment (TME) impacts the therapeutic efficacy of immune checkpoint inhibitors (ICIs). No liquid biomarkers are available to evaluate TME heterogeneity. Here, we investigated the clinical significance of PD-1-binding soluble PD-L1 (bsPD-L1) in gastric cancer (GC) patients and non-small cell lung cancer (NSCLC) patients treated with PD-1/PD-L1 blockade.

**Experimental Design:** We examined bsPD-L1, matrix metalloproteinases (MMPs), and IFN-γ levels in plasma samples from GC patients (n =117) prior to surgery and NSCLC patients (n =72) prior to and two months after ICI treatment and extracellular matrix (ECM) integrity, PD-L1 expression, and T cell infiltration in tumor tissues from 25 GC patients.

**Results:** bsPD-L1 was detected in 17/117 GC patients and 16/72 NSCLC patients. bsPD-L1 was strongly and moderately correlated with MMP13 and MMP3, respectively. In GC, bsPD-L1 expression was associated with IFN-γ levels and intra-tumoral T cell infiltration; MMP13 levels were associated with loss of ECM integrity, allowing tumor cells to access blood vessels. MMP3 and MMP13 levels were altered during ICI treatment. Combination of bsPD-L1 and MMP levels identified two patient groups; bsPD-L1^+^MMP13^high^ in GC and bsPD-L1^+^(MMP3 and MMP13)^increased^ in NSCLC were associated with poor prognosis, and bsPD-L1^+^MMP13^low^ in GC and bsPD-L1^+^(MMP3 or MMP13)^decreased^ in NSCLC were associated with favorable prognosis.

**Conclusions:** Plasma bsPD-L1 and MMP13 levels indicate T cell response and loss of ECM integrity, respectively, in the TME. The combination of bsPD-L1 and MMPs may represent a non-invasive tool to predict recurrence in GC and the efficacy of ICIs in NSCLC.

## Introduction

Immune checkpoint inhibitors (ICIs) targeting programmed cell death 1 (PD-1) and its ligand PD-L1 have revolutionized the treatment of various solid tumors, including non-small cell lung cancer (NSCLC) and gastric cancer (GC) (1–4). However, because of the heterogeneity of the immune microenvironment among patients, only a small subset of patients respond to ICI treatment (5,6). Currently, there are no liquid biomarkers to evaluate the heterogeneity of the tumor microenvironment (TME). Additionally, growing evidence indicates that a significant subset of patients may exhibit hyperprogressive disease (HPD) during immunotherapy (7). HPD is characterized by rapid tumor growth rate and accelerated disease progression following ICI treatment, and is associated with poor prognosis in multiple solid tumor types. However, there are no predictive liquid biomarkers for HPD.

Our early studies using PD-1-deficient mice and blocking antibodies revealed the immunoinhibitory role of PD-1/PD-L1 signaling in anti-tumor and anti-viral immunity (3,8–11). PD-L1 is expressed on various types of cells such as immune cells and tumor cells, and its binding to PD-1 on T cells leads to immunosuppression (3,8). Although tumor PD-L1 expression was the first identified marker to select patients for cancer immunotherapy (12,13), it requires highly invasive tumor biopsies. Besides membrane-bound PD-L1, a soluble form of PD-L1 (sPD-L1) was detected in peripheral blood (14,15) and several mechanisms of sPD-L1 production (e.g., proteolytic cleavage, splicing, and exosomal PD-L1 secretion) have been described (15–19). However, the source and function of sPD-L1 remain uncertain, with many conflicting reports (19–22). Recently, it was reported that PD-L1 is selectively cleaved by matrix metalloproteinases (MMPs), such as MMP13 and MMP9, in vitro (16). MMPs cleave not only extracellular matrix (ECM) components but also non-matrix substrates such as cytokines and membrane proteins (23,24). Although the role of MMPs in cancer invasion and metastasis is well established, little is known about their role in T cell response within the TME.

Based on the hypothesis that functional sPD-L1 binds to the PD-1 receptor, we previously developed an ELISA system to specifically detect PD-1-binding sPD-L1 (bsPD-L1) (25). In this study, we investigated circulating bsPD-L1 and MMP levels and their clinical significance in GC patients as well as NSCLC patients treated with PD-1/PD-L1 blockade.

## Materials and Methods

### Patients and specimens

This study included 117 patients diagnosed with GC between 2017 and 2020 at the Department of Gastrointestinal and Hepato-Biliary-Pancreatic Surgery in Nippon Medical School Hospital, Japan. Blood samples were collected from all patients before surgery. Surgical resection specimens were obtained from 25 GC patients.

The study also included 72 patients diagnosed with NSCLC between 2017 and 2019 at the Department of Pulmonary Medicine and Oncology in Nippon Medical School Hospital, Japan. Blood was collected from patients prior to and at two months after the initiation of checkpoint immunotherapy (nivolumab, n = 20; pembrolizumab, n = 28; atezolizumab, n = 16; durvalumab, n=8). Nivolumab (3 mg/kg, every 2 weeks), pembrolizumab (200 mg/body, every 3 weeks), or atezolizumab (1200 mg/body, every 3 weeks) was administered until disease progression or unacceptable adverse events in a clinical setting. Durvalumab (10 mg/kg, every 2 weeks) was administered after curative chemoradiotherapy. RECIST v1.1 was used to assess efficacy of immunotherapy. Tumor tissue samples from biopsies were obtained from 59 NSCLC patients prior to the initiation of immunotherapy.

Baseline clinical and demographic data were collected from the patients’ medical records. The study protocols (B-2019-005, 28-09-646) were reviewed and approved by the Ethics Committee of Nippon Medical School. All participants provided written informed consent. This study was conducted in accordance with the Declaration of Helsinki.

### Enzyme-linked immunosorbent assay (ELISA)

Human bsPD-L1 levels were measured as previously described (25). MMP3, MMP9, MMP13, and interferon (IFN)-γ concentrations in plasma were determined using ELISA kits (R&D systems, Minneapolis, MN, USA, #DY513, #DY911, #DY511, and #DY285B, respectively) following the manufacturer’s instructions.

### Histological analysis

Serial sections of formalin-fixed paraffin-embedded tumor tissue were subjected to hematoxylin and eosin (H&E), Elastica Masson-Goldner (EMG), or immunohistochemical staining. GC tissue sections were stained using a PD-L1 immunohistochemistry assay (Agilent Technologies, Santa Clara, CA, USA, #28-8 pharmDx) following the manufacturer’s instruction. PD-L1 expression was quantified using the combined positive score (CPS) defined as the number of PD-L1^+^ tumor cells and immune cells (including lymphocytes and macrophages) divided by the total number of viable tumor cells, multiplied by 100. NSCLC tissue sections were stained using a PD-L1 immunohistochemistry assay (Agilent Technologies, #22C3 pharmDx) following the manufacturer’s instruction. PD-L1 expression was quantified using tumor proportion score (TPS) defined as the number of PD-L1^+^ tumor cells divided by the total number of viable tumor cells, multiplied by 100. Pathologists were blinded to clinical information and the evaluation results of other pathologists.

To identify T and B cells, sections were incubated with anti-CD3 (Abcam, Cambridge, UK, #ab5690) and anti-CD20 (Leica Biosystems, Wetzla, Germany, #NCL-L-CD20-L26) antibodies and treated with Histofine Simple Stain MAX-PO (R) and MAX-PO (M) reagents (Nichirei Bioscience, Tokyo, Japan, #424141 and #424131, respectively). Peroxidase activity was visualized using diaminobenzidine. Sections were counterstained with hematoxylin.

Histological images were acquired using a virtual slide scanner (Hamamatsu Photonics, Shizuoka, Japan, #NanoZoomer-SQ, C13140-D03,). The areas of tumor and T cell clusters were calculated using the NDP view2 software (Hamamatsu Photonics). The percentage of T cell cluster area was calculated by dividing the total area of T cell clusters by the total tumor area, multiplied by 100. The numbers of CD3^+^ and CD3^-^ nuclei in different areas of the tumor were counted using the PatholoCount software Ver 1.2.3 (Mitani Corporation, Tokyo, Japan). The percentage of CD3^+^ T cells was calculated by dividing CD3^+^ T cells by the total viable cells, multiplied by 100.

### Statistical analysis

Mann–Whitney U test or Student’s t-test was used to compare numerical variables, and the Pearson’s chi-squared test was used to compare categorical variables. Logistic regression analysis was used to identify factors for bsPD-L1 expression. Receiver operator characteristic (ROC) curve and area under the ROC curve (AUC) were used to assess the discriminatory ability of numerical variables (e.g., MMP3, MMP9, and MMP13 levels). Youden’s index was used to identify the optimal cutoff values. The optimal cutoff value for MMP13 level was used to segregate bsPD-L1^+^ GC patients into two subgroups. Disease-free survival (DFS) was defined as the time from surgery to relapse, second cancer, or all-cause death, whichever came first. Progression-free survival (PFS) was defined as the time from the initiation of treatment to disease progression or death from any cause. Overall survival (OS) was defined as the time from surgery or initiation of treatment to the date of last follow-up or death from any cause. DFS, PFS, and OS were estimated using the Kaplan–Meier method; differences between groups were assessed using the log-rank test. Univariate and multivariate analyses for DFS, PFS, and OS were performed using Cox regression models. Variables associated with prognosis in the univariate analysis were included in the multivariate Cox regression analysis. All tests were two-tailed, and p-values < 0.05 were considered statistically significant. Statistical analyses were performed using JMP software, version 13 (SAS Institute, Cary, NC, USA) and Prism software, version 8 (GraphPad, San Diego, CA, USA).

## Results

### Correlation between bsPD-L1 and MMP13 levels

We investigated the relationship between circulating bsPD-L1 and MMP levels in GC patients (Figure 1A). bsPD-L1 was detected in 17 (14.5%) of 117 GC patients. The expression pattern of bsPD-L1 was similar to that of MMP13, but different from that of MMP9. We found a strong correlation between bsPD-L1 and MMP13 levels and a moderate correlation between bsPD-L1 and MMP3 levels (correlation coefficient [r] = 0.742, p < 0.0001; r = 0.534, p < 0.0001, respectively) (Figure 1B). There was no correlation between bsPD-L1 and MMP9 levels.

**Figure 1.**
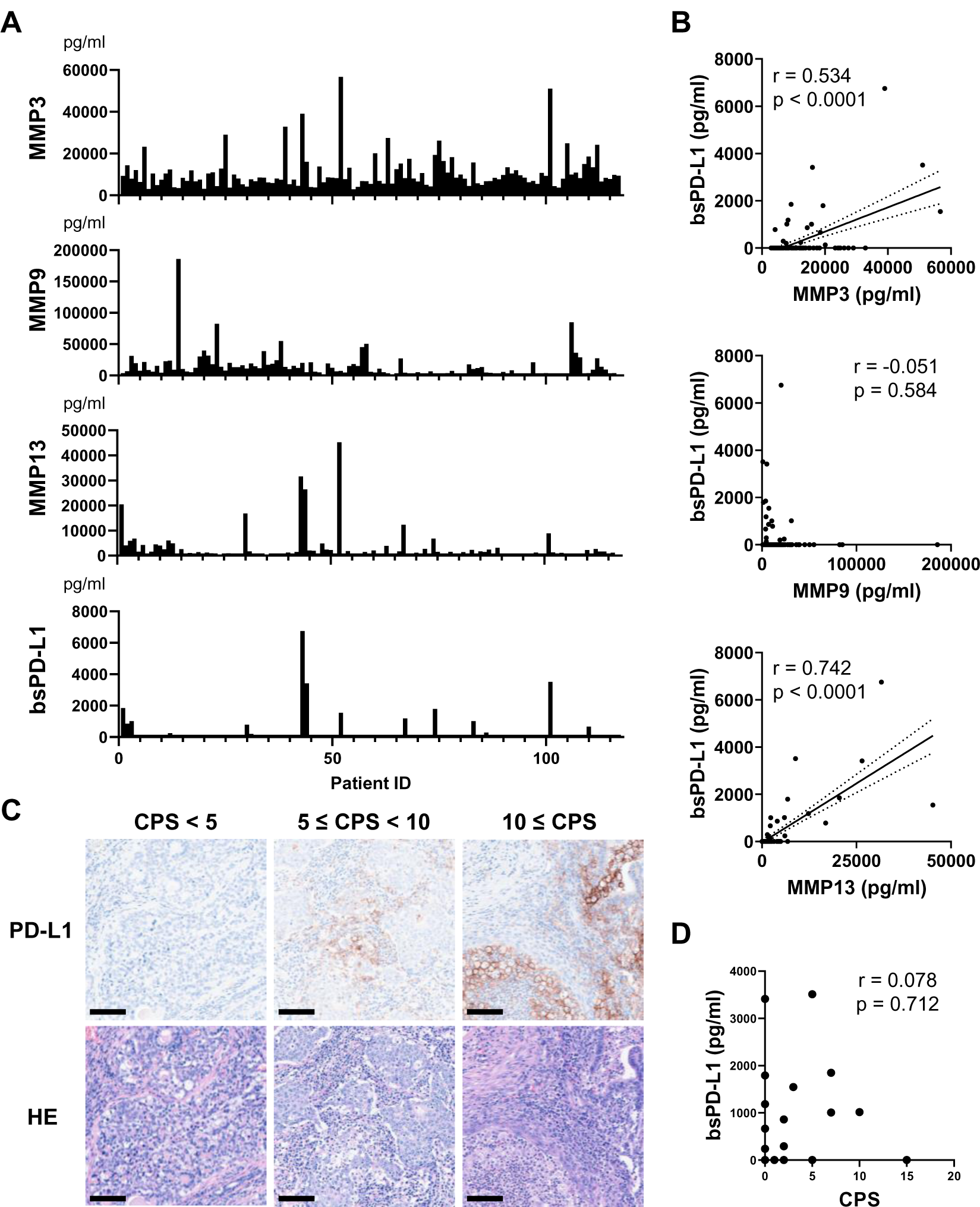
Detection of MMPs and bsPD-L1 in the plasma of GC patients. (A) Plasma MMP and bsPD-L1 levels in 117 GC patients. (B) Correlation between bsPD-L1 and MMP levels in GC patients (n = 117). r indicates the correlation coefficient. (C) Representative images of H&E and anti-PD-L1 immunohistochemical staining in tumor tissues from GC patients with low (CPS < 5), moderate (5 ≤ CPS < 10), and high (CPS ≥ 10) PD-L1 expression. Original magnification, 20×. Scale bars indicate 100 µm. Nuclei were counterstained with hematoxylin (blue). (D) Correlation between bsPD-L1 level and CPS in GC patients (n = 25). r indicates the correlation coefficient.

### Discrepancy between bsPD-L1 levels and tumor PD-L1 expression

We investigated the relationship between plasma bsPD-L1 levels and PD-L1 expression in tumor specimens from 25 GC patients. Tumor PD-L1 expression was quantified by determining the CPS (Figure 1C). We detected low (CPS < 5), moderate (5 ≤ CPS < 10), and high (CPS ≥ 10) PD-L1 expression in tumor specimens of 17, 6, and 2 patients, respectively. There was no significant correlation between bsPD-L1 levels and CPS (Figure 1D), suggesting that plasma bsPD-L1 levels are not associated with tumor PD-L1 expression in GC.

### Correlation between bsPD-L1 and IFN-γ levels

We examined the association between bsPD-L1 levels and clinicopathological features of GC patients. No significant differences were found between the bsPD-L1^+^ and bsPD-L1^-^ groups for any of the variables (Supplementary Table 1). Next, we compared the levels of inflammatory markers between the bsPD-L1^+^ and bsPD-L1^-^ groups (Figure 2). The neutrophil-to-lymphocyte ratio (NLR) and C-reactive protein (CRP) levels tended to be lower in the bsPD-L1^+^ group than in the bsPD-L1^-^ group but without statistical significance. IFN-γ concentration was significantly higher in the bsPD-L1^+^ group than in the bsPD-L1^-^ group (p = 0.0001). Consistent with the results in Figure 1B, the bsPD-L1^+^ group of GC patients had higher levels of MMP3 and MMP13 (but not MMP9) than the bsPD-L1^-^ group.

**Figure 2.**
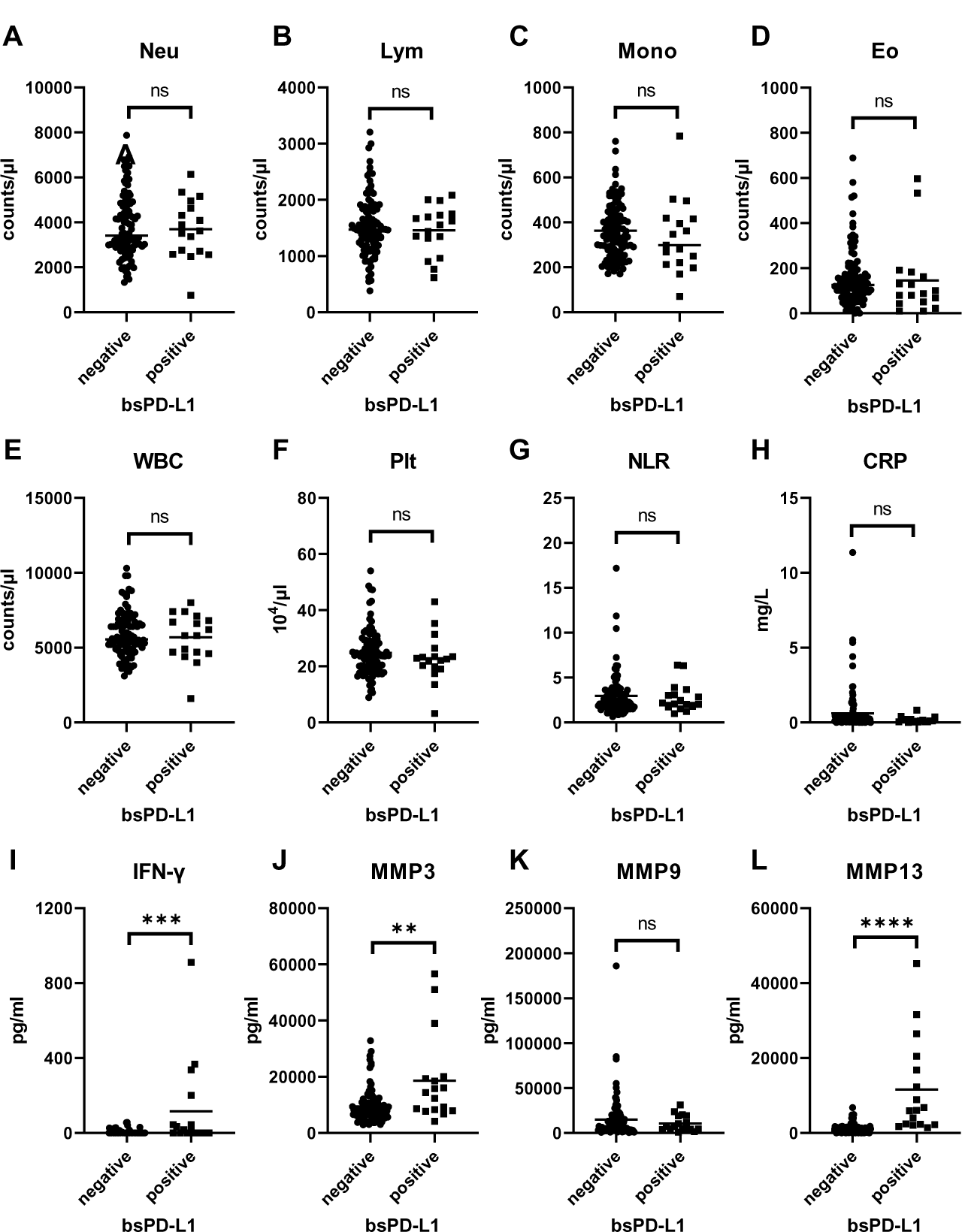
Comparison of inflammatory markers between bsPD-L1^+^ and bsPD-L1^-^ GC patients. Counts of (A) Neutrophil, (B) lymphocyte, (C) monocyte, (D)eosinophil, (E) white blood cell, and (F) platelet, (G) neutrophil-to-lymphocyte ratio, and levels of (H) C-reactive protein, (I) IFN-γ, and (J–L) MMPs in bsPD-L1^+^ (n = 17) and bsPD-L1^-^ (n = 100) GC patients. The horizontal lines indicate the mean. Statistical significance was calculated using the Student’s t-test (A, B, and E) or the Mann–Whitney U test (C, D, F, G, H, I, J, K, and L). ∗∗p < 0.01; ∗∗∗p < 0.001; ∗∗∗∗p < 0.0001; ns, not significant.

### Correlation between bsPD-L1 levels and intra-tumoral T cell infiltration

We investigated the association between bsPD-L1 levels and tumor-infiltrating lymphocytes in tumor tissues from 25 GC patients. Higher numbers of tumor-infiltrating lymphocytes were detected in tumor tissues of bsPD-L1^+^ patients than in those of bsPD-L1^-^ patients (Supplementary Figure 1). Immunohistochemical analysis revealed that the numbers of CD3^+^ T cells were also significantly higher in tumor tissues from bsPD-L1^+^ patients (Supplementary Figure 1B, Figure 3A, and 3B), suggesting that bsPD-L1 levels might reflect T cell infiltration in the TME.

**Figure 3.**
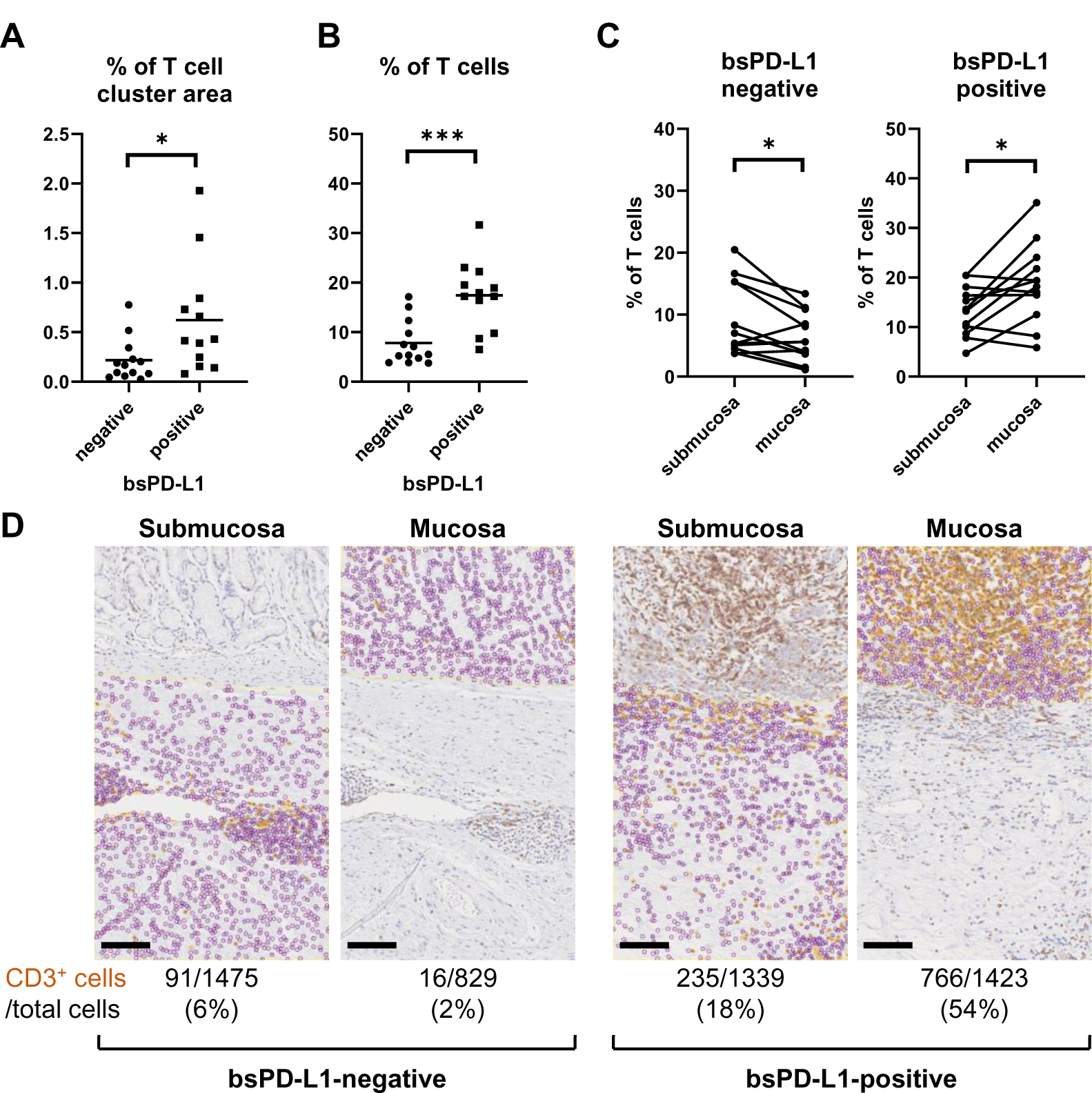
bsPD-L1^+^ GC patients had high numbers of tumor-infiltrating T lymphocytes. (A) The percentages of T cell cluster area within the tumor area and (B) the percentages of CD3^+^ T cells within the total viable cell population in bsPD-L1^+^ (n = 12) and bsPD-L1^-^ (n = 13) GC patients. (C) The percentage of CD3^+^ T cells within the total viable cell population between mucosal and submucosal layers. (D) Representative images of tumor tissues from bsPD-L1^+^ and bsPD-L1^-^ GC patients. Original magnification, 20×. Scale bars indicate 100 µm. Yellow and purple circles indicate CD3^+^ and CD3^-^ nuclei, respectively. Statistical significance was calculated using the unpaired (A and B) or paired (C) Student’s t-test. ∗p < 0.05; ∗∗∗∗p < 0.0001.

Since bsPD-L1^+^ patients had higher levels of the collagenase MMP13 than bsPD-L1^-^ patients (Figure 2L), we evaluated ECM integrity in tumor tissues by EMG staining (Supplementary Figure 1). We found that the layered architecture of mucosal tissue was destroyed in the tumors of bsPD-L1^+^ patients. Specifically, we observed that while the collagen fibers were strongly stained in tumor samples from bsPD-L1^-^ patients, they were only weakly stained in those from bsPD-L1^+^ patients; this was particularly evident in the perivascular area of the tumor, suggesting that collagen fibers surrounding blood vessels were being degraded in the tumors of bsPD-L1^+^ patients.

We further investigated whether the loss of ECM integrity affected T cell localization within the TME (Figure 3C and 3D). Based on the analysis of H&E, EMG, and immunohistochemistry images, we defined the boundary between the mucosal and submucosal layers (Supplementary Figure 1B) and counted the number of CD3^+^ T cells in each area (Figure 3C and 3D). We found a substantial influx of T cells into the mucosal layer (located adjacent to tumor cells) of tumors from bsPD-L1^+^ patients. In contrast, most T cells were localized to the submucosal layer and did not migrate into the mucosal layer of tumors from bsPD-L1^-^ patients (Figure 3C and 3D). These data suggest that bsPD-L1 levels might reflect not only the magnitude of T cell response but also the change in T cell localization in the TME following the loss of ECM integrity.

### Identification of GC patients with a high risk of relapse

Among the bsPD-L1^+^ patients, one notable case (Patient 86) exhibited relatively low levels of MMP13 (Supplementary Figure 2). The patient had a large T cell infiltration in the tumor tissue; however, these T cells were localized in a compartment surrounded by collagen fibers, adjacent to the compartment containing tumor cells. These results suggest that bsPD-L1 and MMP13 may play distinct roles in the TME: bsPD-L1 may regulate T cell response, whereas MMP13 may regulate T cell migration by altering ECM integrity.

We further investigated whether MMP13 levels might affect the migration of tumor cells in the TME (Figure 4A). Histological analysis of tumor tissues from bsPD-L1^+^MMP13^high^ patients revealed the almost complete disappearance of collagen fibers, especially in the perivascular area; this caused tumor cells to localize adjacent to the bloodstream. By contrast, in the tumor tissues from bsPD-L1^+^MMP13^low^ patients, tumor cells were separated from blood flow by thick layers of collagen fibers. These results raise the possibility that MMP13 levels might affect the vascular invasiveness of tumor cells.

**Figure 4.**
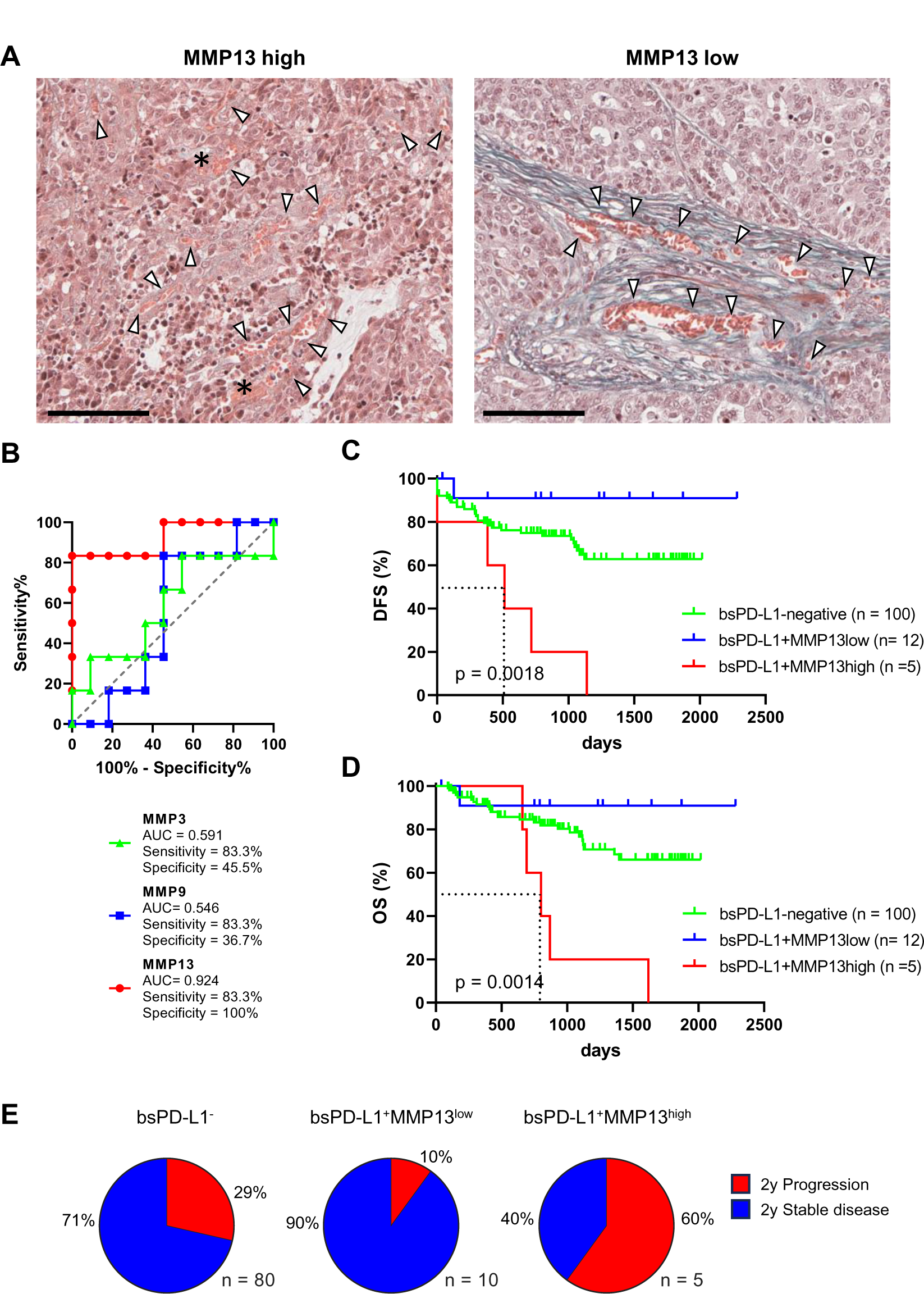
Identification of GC patients with a high risk of relapse. (A) Representative images of EMG staining in tumor tissues from bsPD-L1^+^ GC patients with high or low MMP13 levels, respectively. Original magnification, 20×. Scale bars indicate 100 µm. Collagen fibers and red blood cells are shown in green and orange, respectively. The asterisk and arrow heads represent hemorrhage and blood vessels, respectively. (B) ROC curve analyses of MMP levels to predict DFS in bsPD-L1^+^ GC patients. (C) DFS and (D) OS in bsPD-L1^-^ (n = 100), bsPD-L1^+^MMP13^low^ (n = 12), and bsPD-L1^+^MMP13^high^ (n = 5) GC patients. (E) Pie charts showing the percentages of GC patients with the indicated bsPD-L1 and MMP13 signatures who had progressed at 2 years post-surgery.

The distinct roles of bsPD-L1 and MMP13 in the TME prompted us to investigate whether the combination of bsPD-L1 and MMP13 levels could serve as a biomarker to predict prognosis for GC patients. Patients were first divided into two groups on the basis of bsPD-L1 expression. The bsPD-L1^+^ patients were then subdivided into two groups on the basis of MMP13 levels. To select the optimal cutoff of MMP13 levels for DFS, we performed ROC analysis. The AUC for MMP13 was 0.924, indicating that MMP13 possessed good discriminatory ability compared with MMP3 and MMP9 (0.591 and 0.546, respectively) (Figure 4B).

Patients were categorized using the optimal cutoff of MMP13 levels (16836 pg/ml). The bsPD-L1^+^MMP13^high^ group showed a shorter DFS and OS than the bsPD-L1^+^MMP13^low^ and bsPD-L1^-^ groups (p = 0.0018 and p = 0.0014, respectively; Figure 4C and 4D). The bsPD-L1^+^MMP13^low^ group tended to have a longer DFS and OS than the bsPD-L1^-^ group; however, this did not reach statistical significance. The median DFS of the bsPD-L1^+^MMP13^high^ was 513 days, while those of the bsPD-L1^+^MMP13^low^ and bsPD-L1^-^ groups were not reached during this study. Strikingly, 60% of the bsPD-L1^+^MMP13^high^ patients progressed ∼2 years after surgery (Figure 4E). Moreover, all bsPD-L1^+^MMP13^high^ patients died within 5 years of surgery (Figure 4D). These results suggest that patients with a high risk of recurrence were enriched in the bsPD-L1^+^MMP13^high^ group.

Multivariate analysis using the Cox regression model revealed that bsPD-L1^+^MMP13^high^ status, lymph node metastasis, and distant metastasis were independent poor prognostic factors for DFS in GC patients (p = 0.015, p = 0.003, and p < 0.0001, respectively; Supplementary Table 2). Among these parameters, distant metastasis was the only independent poor prognostic factor for OS (p = 0.001; Supplementary Table 2).

### Change in MMP levels during ICI treatment in NSCLC patients

We next investigated bsPD-L1 and MMP levels in NSCLC patients during ICI treatment (Supplementary Figure 3). At baseline, bsPD-L1 was detected in 16 out of 72 NSCLC patients (22.2%). Consistent with observations in GC patients, the expression pattern of bsPD-L1 was similar to that of MMP13, but different from that of MMP9 (Supplementary Figure 3A). bsPD-L1 was strongly and moderately correlated with MMP13 (r = 0.821, p < 0.0001) and MMP3 (r = 0.372, p = 0.0013), respectively (Supplementary Figure 3B). During ICI treatment, bsPD-L1 and MMPs showed different kinetics changes (Supplementary Figure 3C). At two months of treatment, 44, 38, and 15 (61%, 53%, and 21%) of the 72 patients showed increased MMP3, MMP9, and MMP13 levels, respectively, while 28, 34, and 20 patients (39%, 47%, and 28%) showed decreased MMP3, MMP9, and MMP13 levels, respectively. bsPD-L1 levels were increased in 12 patients (17%), whereas only one patient (1.3%) showed a negligible decrease in bsPD-L1.

### High MMP13 was an independent factor associated with bsPD-L1 expression

To identify factors associated with bsPD-L1 expression, NSCLC patients were divided into two groups on the basis of bsPD-L1 expression. Univariate analysis revealed that the bsPD-L1^+^ group contained fewer patients with a smoking history than the bsPD-L1^-^ group (p = 0.0441, Supplementary Table 3). There was no significant correlation between bsPD-L1 levels and tumor PD-L1 expression determined by TPS (p = 0.1724). We then compared the levels of inflammatory markers between the bsPD-L1^+^ and bsPD-L1^-^ groups (Supplementary Figure 4). Neutrophil count and CRP levels were significantly lower in the bsPD-L1^+^ group than in the bsPD-L1^-^ group (p = 0.0433, p = 0.0260, respectively). Among the MMPs, only MMP13 levels were significantly higher in the bsPD-L1^+^ group than in the bsPD-L1^-^ group (p < 0.0001). Multivariate analysis demonstrated that high MMP13 was an independent factor associated with bsPD-L1 expression (p = 0.0006, Supplementary Table 4).

### Identification of NSCLC patients without benefit from ICI treatment

To investigate the clinical significance of bsPD-L1 in NSCLC patients treated with ICIs, we evaluated the association between bsPD-L1 expression and PFS or OS. We noted a crossing of the OS curves at day 700 (Figure 5A). Before day 700, the OS rate was lower in bsPD-L1^+^ patients than in bsPD-L1^-^ patients, suggesting an enrichment of patients with rapid progression in the bsPD-L1^+^ group. The OS curve of bsPD-L1^+^ patients reached a plateau at day 400, and the OS rate became higher in the bsPD-L1^+^ patients than in the bsPD-L1^-^ patients after day 700, suggesting an enrichment of long-term responders in the bsPD-L1^+^ group. These results raise the possibility that patients with HPD and with durable clinical benefit from ICIs were enriched in the bsPD-L1^+^ groups.

**Figure 5.**
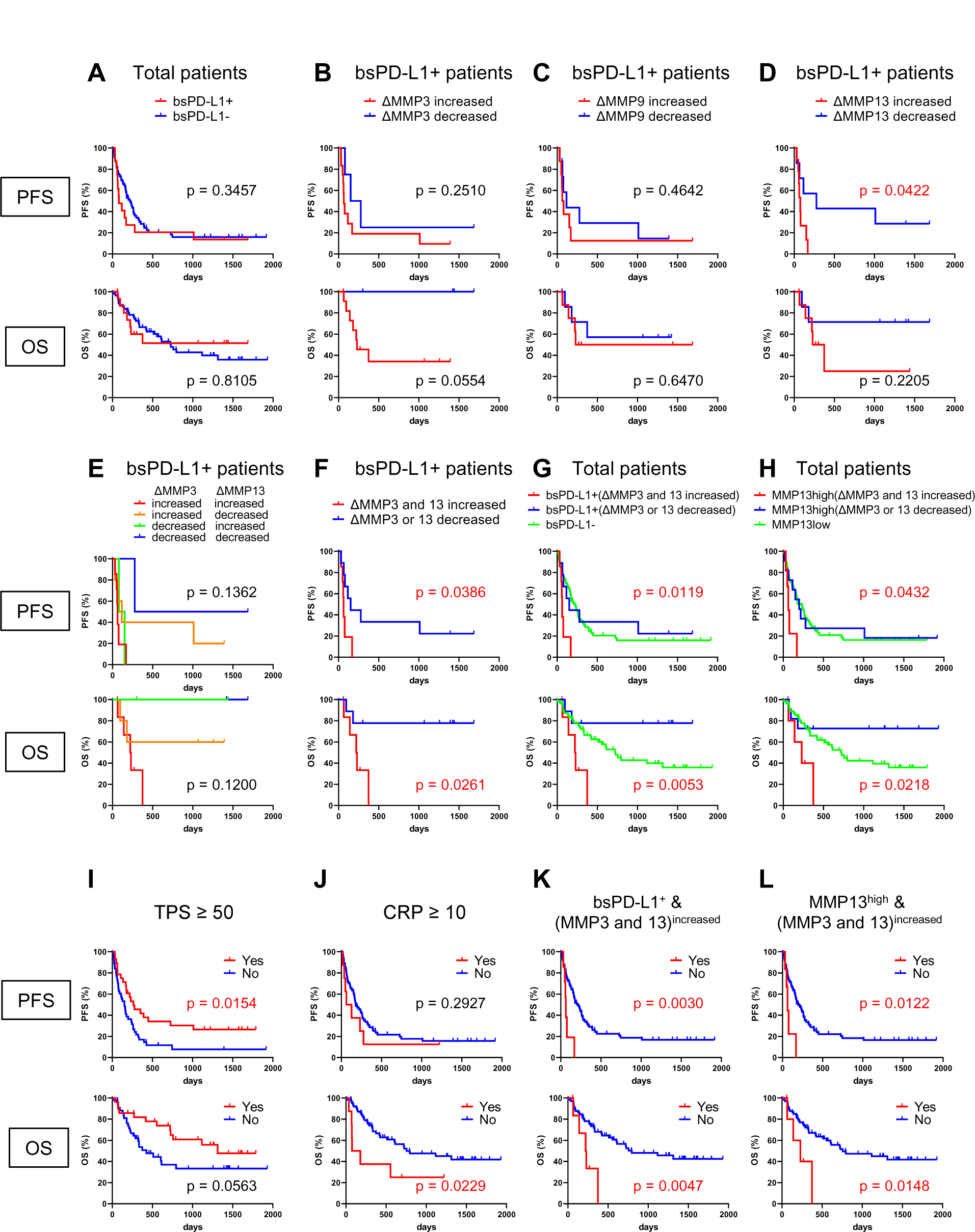
Identification of NSCLC patients without benefit from ICI treatment. (A) PFS and OS of bsPD-L1^+^ (n = 16) and bsPD-L1^-^ (n = 56) NSCLC patients. (B) PFS and OS of MMP3^increased^ (n = 4) and MMP3^decreased^ (n = 12) bsPD-L1^+^ patients. (C) PFS and OS of MMP9^increased^ (n = 8) and MMP9^decreased^ (n = 8) bsPD-L1^+^ patients. (D) PFS and OS of MMP13^increased^ (n = 7) and MMP13^decreased^ (n = 9) bsPD-L1^+^ patients. (E) PFS and OS of _MMP3_increased_MMP13_increased _(n = 7), MMP3_increased_MMP13_decreased _(n = 5),_ MMP3^decreased^MMP13^increased^ (n = 2), and MMP3^decreased^MMP13^decreased^ (n = 2) bsPD-L1^+^ patients. (F) PFS and OS of (MMP3 and MMP13)^increased^ (n = 7) and (MMP3 or MMP13)^decreased^ (n = 9) bsPD-L1^+^ patients. (G) PFS and OS of bsPD-L1^-^ (n = 56), bsPD-L1^+^(MMP3 and MMP13)^increased^ (n = 7), and bsPD-L1^+^(MMP3 or MMP13)^decreased^ (n = 9) NSCLC patients. (H) PFS and OS of MMP13^low^ (n = 55), MMP13^high^(MMP3 and MMP13)^increased^ (n = 6), and MMP13^high^(MMP3 or MMP13)^decreased^ (n = 11) NSCLC patients. (I) PFS and OS of NSCLC patients stratified by TPS (≥ 50, n = 28 vs. < 50, n = 44). (J) PFS and OS of NSCLC patients stratified by CRP level (≥ 10 mg/L, n = 8 vs. < 10 mg/L, n = 64). (K) PFS and OS of NSCLC patients stratified by bsPD-L1^+^(MMP3 and MMP13)^increased^ status (yes, n = 7 vs. no, n = 65). (L) PFS and OS of NSCLC patients stratified by MMP13^high^(MMP3 and MMP13)^increased^ status (yes, n = 6 vs. no, n = 66).

Since MMP levels altered during immunotherapy (Supplementary Figure 3C), we investigated whether the change in MMP levels could stratify responders from non-responders among bsPD-L1^+^ patients. bsPD-L1^+^ patients were subdivided into two groups on the basis of change in MMP levels at 2 months after initiation of immunotherapy (Figure 5B–D). bsPD-L1^+^MMP3^increased^ patients had a trend toward shorter OS than bsPD-L1^+^MMP3^decreased^ patients (p = 0.0554). bsPD-L1^+^MMP13^increased^ patients showed a shorter PFS than bsPD-L1^+^MMP13^decreased^ patients (p = 0.0422). We found no association between MMP9 change and either PFS or OS (p = 0.4642, p = 0.6470, respectively). These results suggest that the increase of MMP13 or MMP3 was associated with poor clinical outcomes in bsPD-L1^+^ patients. Next, to increase predictive accuracy, we evaluated the combination of MMP3 and MMP13 change (Figure 5E and 5F). bsPD-L1^+^ patients were categorized on the basis of MMP changes: _(i) MMP3_increased_MMP13_increased_, (ii) MMP3_increased_MMP13_decreased_, (iii) MMP3_decreased_MMP13_increased, and (iv) MMP3^decreased^MMP13^decreased^. MMP13 increase was strongly associated with a high risk of progression, whereas MMP3 increase was strongly associated with a high risk of death, suggesting that MMP3 and MMP13 may play distinct roles in the TME (Figure 5E). These analyses demonstrate that the increase of both MMP3 and MMP13 may identify patients without benefit from ICI (Figure 5F).

Finally, we evaluated the clinical potential of combining bsPD-L1 with MMP change (Figure 5G). NSCLC patients were grouped on the basis of baseline bsPD-L1 expression, and bsPD-L1^+^ patients were subdivided by MMP3 and MMP13 change. bsPD-L1^+^(MMP3 and MMP13)^increased^ patients showed a shorter PFS and OS than other groups (p = 0.0119, p = 0.0053, respectively). bsPD-L1^+^(MMP3 or MMP13)^decreased^ patients tended to have a longer OS than the bsPD-L1^-^ group, but without significance. The median PFS of bsPD-L1^+^(MMP3 and MMP13)^increased^, bsPD-L1^+^(MMP3 or MMP13)^decreased^, and bsPD-L1^-^ groups was 63, 151, and 217 days, respectively. The median OS of bsPD-L1^+^(MMP3 and MMP13)^increased^, bsPD-L1^+^(MMP3 or MMP13)^decreased^, and bsPD-L1^-^ groups were 225 days, not reached, and 726 days, respectively.

To reduce variables in the combined analysis, we investigated whether baseline bsPD-L1 could be replaced with baseline MMP13 (Figure 5H), because bsPD-L1 strongly correlated with MMP13 (Supplementary Figure 3B). To select the optimal cutoff of MMP13 levels for bsPD-L1 expression, we performed ROC analysis. The AUC was 0.950, demonstrating a good discriminatory ability. NSCLC patients were divided into MMP13-high and low groups using the cutoff of baseline MMP13 (985 pg/ml); MMP13^high^ patients were subdivided by MMP3 and MMP13 change. The MMP13^high^(MMP3 and MMP13)^increased^ patients showed a shorter PFS and OS than other groups (p = 0.0432, p = 0.0218, respectively). The MMP13^high^(MMP3 or MMP13)^decreased^ patients tended to have a longer OS than MMP13^low^ group but without significance. The median PFS of MMP13^high^(MMP3 and MMP13)^increased^, MMP13^high^(MMP3 or MMP13)^decreased^, and MMP13^low^ group groups was 63, 198, and 232 days, respectively. The median OS of MMP13^high^(MMP3 and MMP13)^increased^, MMP13^high^(MMP3 or MMP13)^decreased^, and MMP13^low^ groups was 229 days, not reached, and 719 days, respectively.

Multivariate analysis using the Cox regression model revealed that bsPD-L1^+^(MMP3 and MMP13)^increased^ status was an independent poor prognostic factor for both PFS and OS in NSCLC patients (p = 0.0096 and p = 0.0166, respectively; Table 1). Low tumor PD-L1 expression (TPS < 50) was an independent poor factor for PFS (p = 0.0069), but not for OS. Age (≥ 65) and high CRP levels (≥ 10 mg/L) were independent poor factors for OS (p = 0.0274 and p = 0.0160, respectively), but not for PFS. MMP13^high^(MMP3 and MMP13)^increased^ status was also an independent poor prognostic factor for both PFS and OS among NSCLC patients (p = 0.0219 and p = 0.0332, respectively; Supplementary Table 5). Compared with TPS and CRP, both bsPD-L1^+^(MMP3 and MMP13)^increased^ and MMP13^high^(MMP3 and MMP13)^increased^ status had high predictive accuracy to identify patients with a high risk of disease progression and death (Figure 5I–L).

**Table 1.** Univariate and multivariate analysis for PFS and OS in NSCLC patients treated with PD-1/PD-L1 blockade.

| Variables | Univariate analysis for PFS |  | Multivariate analysis for PFS |  |
| --- | --- | --- | --- | --- |
|  | HR (95% CI) | p value | HR (95% CI) | p value |
| Age ≥ 65 (vs. < 65) | 1.07 (0.64 - 1.83) | 0.793 | | |
| Male (vs. Female) | 0.96 (0.54 - 1.86) | 0.910 |  |  |
| Lymph node metastasis (yes vs. no) | 1.15 (0.59 - 2.50) | 0.704 |  |  |
| Distant metastasis (yes vs. no) | 0.72 (0.39 - 1.26) | 0.251 |  |  |
| CRP ≥ 10 (vs. < 10) | 1.52 (0.63 - 3.13) | 0.329 | | |
| NLR ≥ 3 (vs. < 3) | 0.95 (0.56 - 1.60) | 0.846 | | |
| TPS < 50 (vs. ≥ 50) | 2.35 (1.30 - 4.32) | 0.0045 | 2.27 (1.25 - 4.21) | 0.0069 |
| bsPD-L1<sup>+</sup>(MMP3 and MMP13) <sup>increased</sup> (yes vs.no) | 3.57 (1.31 - 8.32) | 0.0157 | 4.16 (1.47 - 10.31) | 0.0096 |

| Variables | Univariate analysis for OS |  | Multivariate analysis for OS |  |
| --- | --- | --- | --- | --- |
|  | HR (95% CI) | p value | HR (95% CI) | p value |
| Age ≥ 65 (vs. < 65) | 2.07 (1.05 - 4.28) | 0.0348 | 2.16 (1.09 - 4.53) | 0.0274 |
| Male (vs. Female) | 1.09 (0.52 - 2.57) | 0.821 |  |  |
| Lymph node metastasis (yes vs. no) | 0.72 (0.33 - 1.79) | 0.449 |  |  |
| Distant metastasis (yes vs. no) | 0.56 (0.25 - 1.14) | 0.112 |  |  |
| CRP ≥ 10 (vs. < 10) | 2.67 (1.00 - 6.02) | 0.0500 | 3.56 (1.30 - 8.31) | 0.0160 |
| NLR ≥ 3 (vs. < 3) | 1.25 (0.65 - 2.42) | 0.497 | | |
| TPS < 50 (vs. ≥ 50) | 2.06 (0.99 - 4.45) | 0.0538 | | |
| bsPD-L1<sup>+</sup>(MMP3 and MMP13) <sup>increased</sup> (yes vs.no) | 3.81 (1.25 - 9.60) | 0.0213 | 4.11 (1.34 - 10.57) | 0.0166 |

## Discussion

In this study, we investigated bsPD-L1 and MMP levels and their clinical significance in GC and NSCLC patients. bsPD-L1 was detected in 15% and 22% of GC and NSCLC patients, respectively. bsPD-L1 was strongly and moderately correlated with MMP13 and MMP3, respectively. In GC, bsPD-L1 expression was associated with IFN-γ levels and intra-tumoral T cell infiltration, suggesting that bsPD-L1 might be a good indicator for T cell response in the TME. MMP13 levels were associated with loss of ECM integrity, which may lead to increased T cell infiltration as well as tumor invasion.

To the best of our knowledge, this study is the first to report an association between bsPD-L1 and MMP13 levels in cancer patients (Figure 1B). A previous study showed that PD-L1 is selectively cleaved by MMP13 and MMP9 in vitro (16). Our results showed a strong correlation of bsPD-L1 with MMP13, but not MMP9, in both GC and NSCLC patients, suggesting that MMP13 may be a key enzyme involved in bsPD-L1 production in vivo. MMP13 is secreted as an inactive proenzyme that is activated when cleaved by extracellular proteinases (26,27); therefore it is unlikely that MMP13 is involved in PD-L1 alternative splicing and exosomal PD-L1 secretion.

We established a novel diagnostic method to non-invasively evaluate ECM and identify patients with a high risk of rapid progression using liquid biomarkers for host T cell response and cancer progression (Supplementary Figure 5 and 6). Baseline bsPD-L1 may be a biomarker for host inherent T cell immune status prior to therapeutic intervention. MMP13 and its activator MMP3 may be used as biomarkers for cancer progression. Notably, MMP13 may be involved in both T cell response and tumor invasion via PD-L1 cleavage and degradation of collagen fibers. The balance between host T cell response and cancer progression is important for the clinical outcome of patients. High levels of MMP13 and its activator MMP3 promote cancer progression rather than T cell response. As shown in Figure 5, the combination of bsPD-L1 and MMP status had higher predictive accuracy to identify NSCLC patients without benefit from ICI treatment than tumor PD-L1 expression, which is used in routine diagnostics for patient selection (12,13,28).

The combination of circulating bsPD-L1 and MMPs has great advantages to stratify responders from non-responders, because they are liquid biomarkers and do not require tumor resections or biopsies. While tumor PD-L1 expression, mutation burden, or phenotype of tumor-infiltrating lymphocytes can identify patient subgroups with or without benefit from ICI treatment, these analyses require tumor tissues acquired by highly invasive tumor biopsies (28–32). Thus, circulating bsPD-L1 and MMP13 may represent a robust and non-invasive tool to identify patients without benefit. Furthermore, we demonstrated that the stratification can be simplified by replacing baseline bsPD-L1 with baseline MMP13 (Figure 5H and 5L).

In contrast to previous reports of sPD-L1 in cancer patients, we focused on sPD-L1 with PD-1-binding ability. There are many conflicting reports on the function of sPD-L1 (19–22). The discrepancies may be from qualitative differences in sPD-L1, because not all sPD-L1 binds to its receptor. Thus, bsPD-L1 might be a more suitable marker for the evaluation of T cell response than sPD-L1. We showed that bsPD-L1 levels positively correlated with IFN-γ levels and intra-tumoral T cell infiltration. Our findings raise the possibility that bsPD-L1 might function as a natural endogenous PD-1 blocker. bsPD-L1 may be useful for pretreatment stratification to reduce overtreatment.

We found a discrepancy between plasma bsPD-L1 levels and tumor PD-L1 expression in GC (Figure 1D). Anti-PD-L1 antibodies used for immunohistochemistry recognize the extracellular region of PD-L1 and thus fail to detect cleaved PD-L1 in tumor tissues. Whether bsPD-L1 is released from the tumor or other tissues remains uncertain. Tumor PD-L1 expression is used for pretreatment stratification in routine diagnosis (12,13,28), and thus cleavage of membrane-bound PD-L1 should be taken into consideration. Evaluation of PD-L1 expression using immunohistochemistry alone may lead to the misinterpretation of a patient’s cancer-immune status.

We used bsPD-L1 and MMP13 to categorize GC patients into three groups: (i) bsPD-L1^-^ (immune-silent phenotype); (ii) bsPD-L1^+^MMP13^low^ (immune-activated phenotype with ECM integrity); and (iii) bsPD-L1^+^MMP13^high^ (immune-activated phenotype without ECM integrity). We observed T cell infiltration in both the bsPD-L1^+^MMP13^low^ and bsPD-L1^+^MMP13^high^ groups. However, bsPD-L1^+^MMP13^high^ patients had an increased risk of disease progression, whereas bsPD-L1^+^MMP13^low^ patients had a favorable prognosis. Our results are consistent with the study by Giraldo et al. showing that clear cell renal cell carcinoma with extensive T cell infiltration were subdivided into two groups with a good or poor prognosis (6,33).

We found that MMP13 and MMP3 levels significantly changed during immunotherapy. The combination of baseline bsPD-L1 expression with MMP3 and/or MMP13 change categorized NSCLC patients treated with ICIs into three groups: (i) baseline bsPD-L1^-^, (ii) baseline bsPD-L1^+^(MMP3 and MMP13)^increased^, and (iii) baseline bsPD-L1^+^(MMP3 or MMP13)^decreased^. The bsPD-L1^+^(MMP3 and MMP13)^increased^ patients were associated with poor clinical outcome with rapid progression, whereas the bsPD-L1^+^(MMP3 or MMP13)^decreased^ patients were associated with good clinical outcome with long survival. These results suggest that both patients with HPD and durable clinical benefit were enriched in the bsPD-L1^+^ groups. Typical immunotherapy-induced crossing and stable plateau of survival curves were observed in many trials (34–37). We speculate that the crossing of OS curves, shown in Figure 5A, is most likely from the two populations with poor and good prognosis. Baseline bsPD-L1 in combination with MMP change may be a powerful non-invasive tool to select patients for cancer immunotherapy. MMPs are likely to be more sensitive to environmental factors such as ICIs than bsPD-L1 (Supplementary Figure 3C). Our new method combining bsPD-L1 and MMPs may be applied to predict prognosis for patients receiving treatments other than immunotherapy such as radiotherapy, chemotherapy, and molecular targeted therapy.

In this study, we showed the distinct clinical value of MMP3 and MMP13. MMP13 expression had a significant impact on the architecture of connective tissue and the migration of both T cells and tumor cells in the TME. MMP13 increase was strongly associated with a high risk of progression (Figure 5D and 5E), whereas MMP3 increase was strongly associated with a high risk of death (Figure 5B and 5E). MMP3 is an upstream MMP activator and activates a wide range of MMPs including MMP13 (26,27). Compared with MMP13, MMP3 is abundant and degrades various ECM substrates as well as non-matrix substrates, including cytokines and chemokines (38). The differences in the proteolytic cascade and preferred substrates may explain the distinct roles of MMP3 and MMP13 in the TME; MMP13 may be more specialized for the regulation of ECM integrity, while MMP3 may have broader roles in cancer progression. MMPs degrade ECM components and play key roles in cancer invasion and metastasis (23,24). Over 50 MMP inhibitors have been investigated in clinical trials involving patients with various cancers; however, all of these trials have failed (24). One possible reason for this outcome is the limited understanding of the effect of MMPs on immune checkpoint molecules; while MMP-mediated degradation of ECM components promotes cancer progression, it could conversely promote intra-tumoral T cell infiltration (Figure 3 and Supplementary Figure 1). Additionally, the cleavage of immune checkpoint molecules by MMPs may enhance anti-tumor T cell responses. Collectively, our results shed light on the opposing roles of MMPs in the TME. MMP13 has been reported to cleave and release tumor necrosis factor (TNF)-α (39). Both TNF-α and IFN-γ are secreted by activated T cells and induce PD-L1 expression in tumor tissues, which might serve as a reservoir of bsPD-L1. We noted some patients with extremely high levels of both bsPD-L1 and MMP13, suggesting that MMP13 in concert with bsPD-L1 may generate a positive feedback loop for T cell activation. Thus, we did not remove extreme data as outliers in correlation analyses (Figure 1 and Supplementary Figure 3).

Our results demonstrate that plasma bsPD-L1 and/or MMPs levels may be prognostic liquid biomarkers for predicting the efficacy of immunotherapy. However, our study had several limitations. This was a retrospective study with a relatively small sample size. Blood samples older than 3 years are not suitable for measuring the concentrations of bsPD-L1 and MMPs, since these factors are unstable proteolytic products. The time interval to evaluate the change in MMPs should be shortened for early diagnostics and selection of therapeutic approaches. Larger prospective studies are required to validate our results.

## Data Availability

All data produced in the present work are contained in the manuscript.

## Author contributions

Conceptualization: Y. I.; Methodology: Y. I., F. A., T. F., and R. O.; Investigation: F. A., S. K., T. F., R. T., and Y. M.; Data analysis: Y. I., T. K., F. A., R. T., T. S., T. A., and M. H.; Resources: T. K., F. A., Y. K, H. Y., M. S., and A. G.; Funding acquisition: Y. I.; Supervision: Y. I. and A. G.; Writing – original draft: Y. I., F. A., and T. K.; Writing – review & editing: all authors.

## Acknowledgments

This work was partially supported by the Grant-in-Aid for Scientific Research from the Japan Society for the Promotion of Science (JP19K07783 and 22K07262 to Y. I.) and a research grant from Sysmex Corporation (to Y. I.). We thank K. Nishimaki for technical and secretarial assistance and R. Yabe, R. Okamoto, and N. Mizushima for helpful discussions.

