## Supplementary Fig.1 for "Clinical significance of PD-1-binding soluble PD-L1 and MMPs in the microenvironment of gastric cancer and non-small cell lung cancer treated with PD-1/PD-L1 blockade"

### Supplementary Figure 1

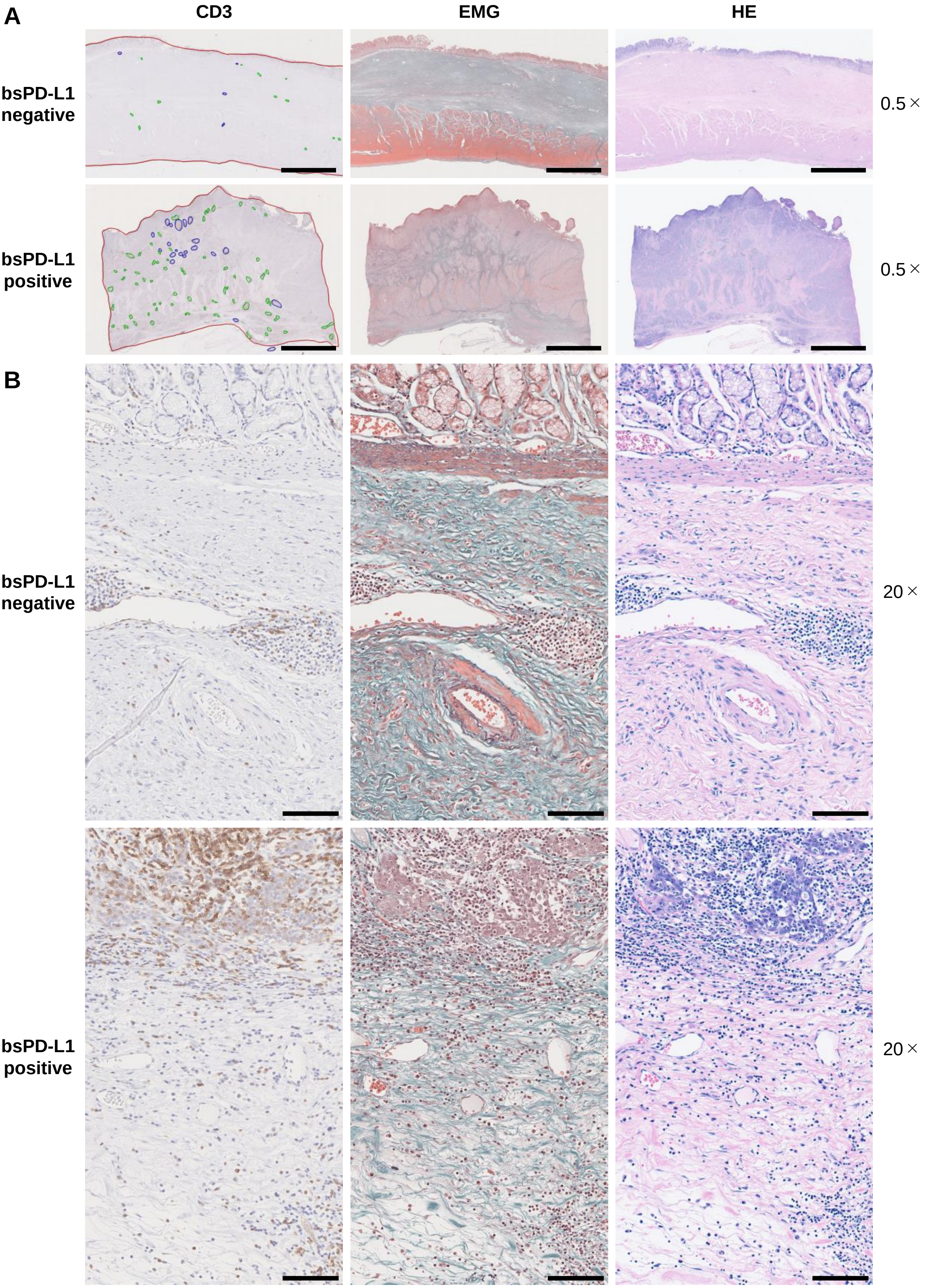


**Histological analysis of T cell infiltration and extracellular matrix integrity in tumor tissues.** Serial tumor tissue sections from bsPD-L1$^{+}$ (n = 12) or bsPD-L1$^{-}$ (n = 13) GC patients were analyzed using H&E, EMG, or anti-CD3 immunohistochemical staining. Representative images at low magnification (0.5×; A) and high magnification (20×; B); scale bars indicate 5 mm (A) and 100 μm (B), respectively. In the immunohistochemical staining images, tumor regions, T cell cluster areas, and B cell follicles are indicated as red, green, and blue lines, respectively. In the EMG staining images, collagen fibers, elastic fibers, red blood cells, and muscle are shown in green, dark purple, orange, and red, respectively.
