## Supplementary Fig.2 for "Clinical significance of PD-1-binding soluble PD-L1 and MMPs in the microenvironment of gastric cancer and non-small cell lung cancer treated with PD-1/PD-L1 blockade"

**Supplementary Figure 2**

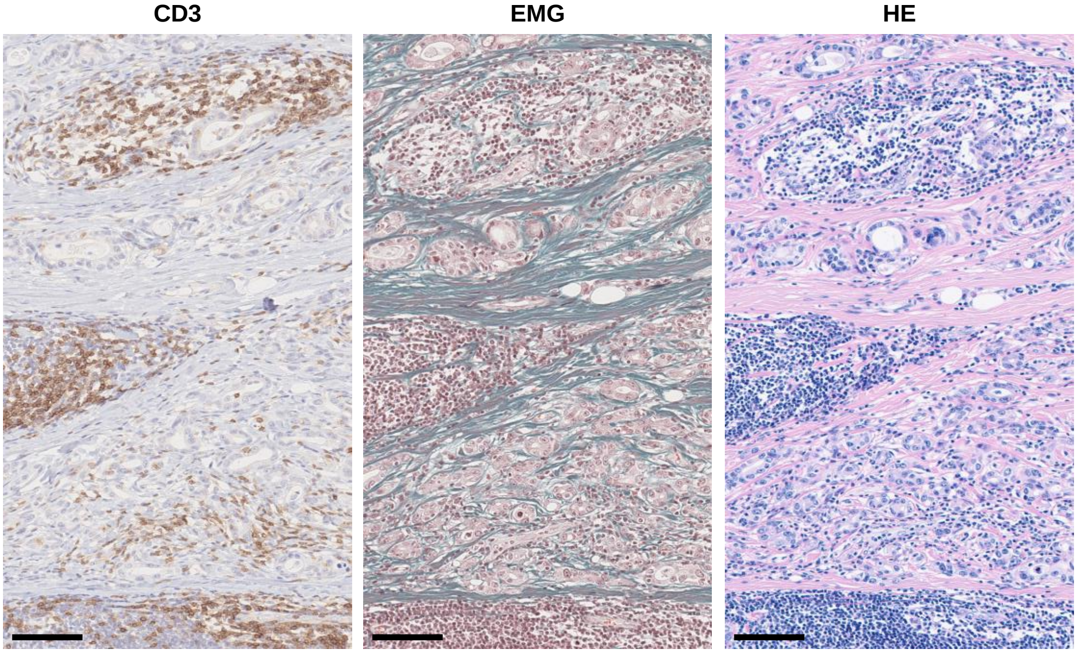


**Histological analysis of gastric cancer tissue from Patient 86.** Serial sections of gastric cancer tissue from Patient 86 were analyzed by H&E, EMG, and anti-CD3 immunohistochemical staining. Original magnification, 20×. Scale bars indicate 100 µm.
