## Supplementary Fig.3 for "Clinical significance of PD-1-binding soluble PD-L1 and MMPs in the microenvironment of gastric cancer and non-small cell lung cancer treated with PD-1/PD-L1 blockade"

### Supplementary Figure 3

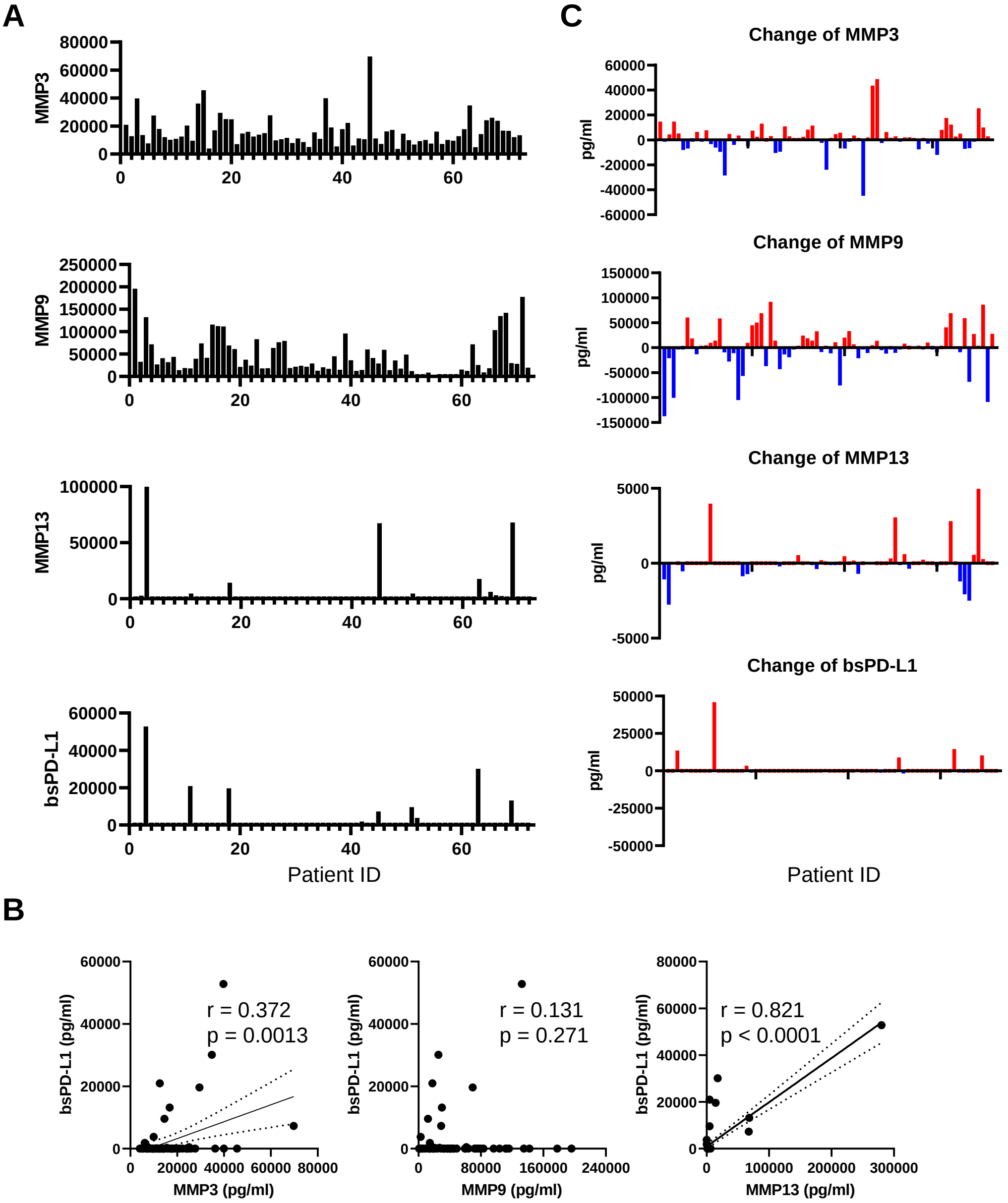


**Detection of MMPs and bsPD-L1 in the plasma of NSCLC patients.**

(A) MMP and bsPD-L1 levels in plasma samples from 72 NSCLC patients. (B) The correlation between the bsPD-L1 and MMP levels in 72 NSCLC patients. r indicates the correlation coefficient. (C) Change of MMP levels in 72 NSCLC patients at 2 months of ICI treatment.
