## Supplementary Fig.4 for "Clinical significance of PD-1-binding soluble PD-L1 and MMPs in the microenvironment of gastric cancer and non-small cell lung cancer treated with PD-1/PD-L1 blockade"

### Supplementary Figure 4

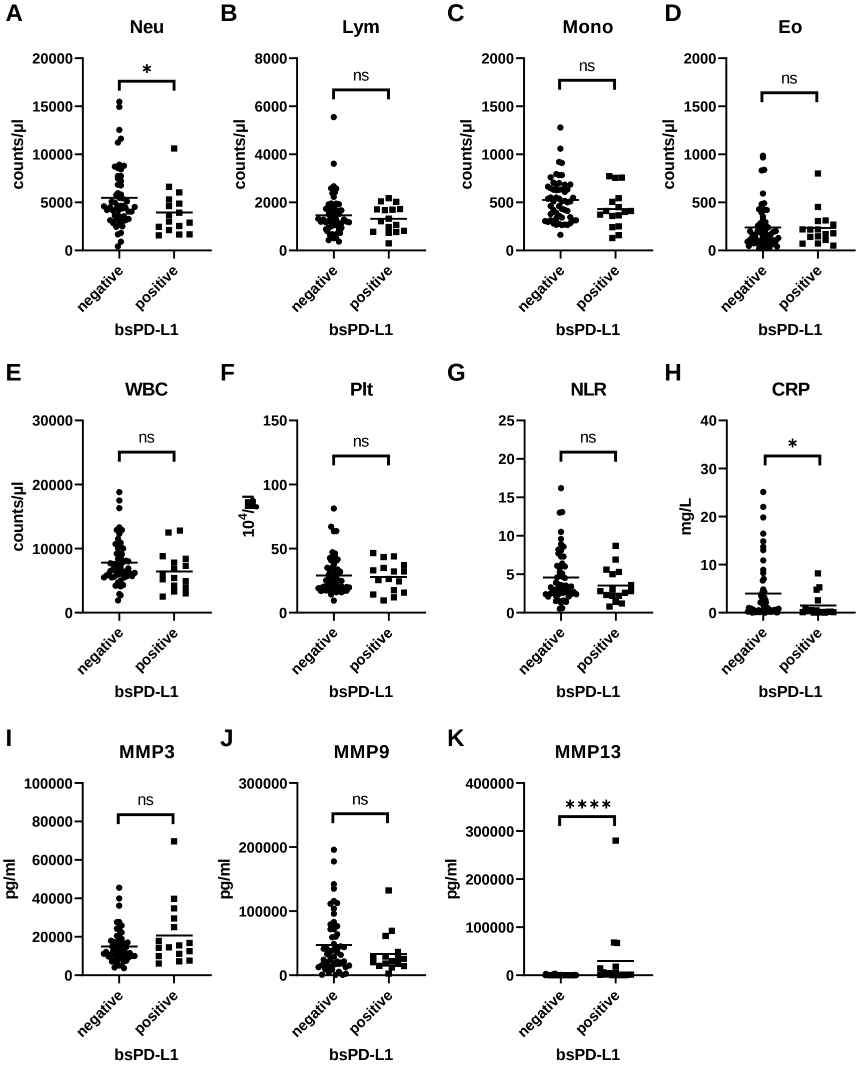


**Comparison of inflammatory markers between bsPD-L1$^+$ and bsPD-L1$^-$ NSCLC patients.** Counts of (A) Neutrophil, (B) lymphocyte, (C) monocyte, (D) eosinophil, (E) white blood cell, and (F) platelet, (G) neutrophil-to-lymphocyte ratio, and levels of (H) C-reactive protein, and (I–K) MMPs in bsPD-L1$^+$ (n = 16) and bsPD-L1$^-$ (n = 56) NSCLC patients. The horizontal lines indicate the mean. Statistical significance was calculated using the Student's t-test (A, B, and E) or the Mann–Whitney U test (C, D, F, G, H, I, J, and K). $*p < 0.05$; $****p < 0.0001$; ns, not significant.
