## Supplementary Fig.5 for "Clinical significance of PD-1-binding soluble PD-L1 and MMPs in the microenvironment of gastric cancer and non-small cell lung cancer treated with PD-1/PD-L1 blockade"

### Supplementary Figure 5

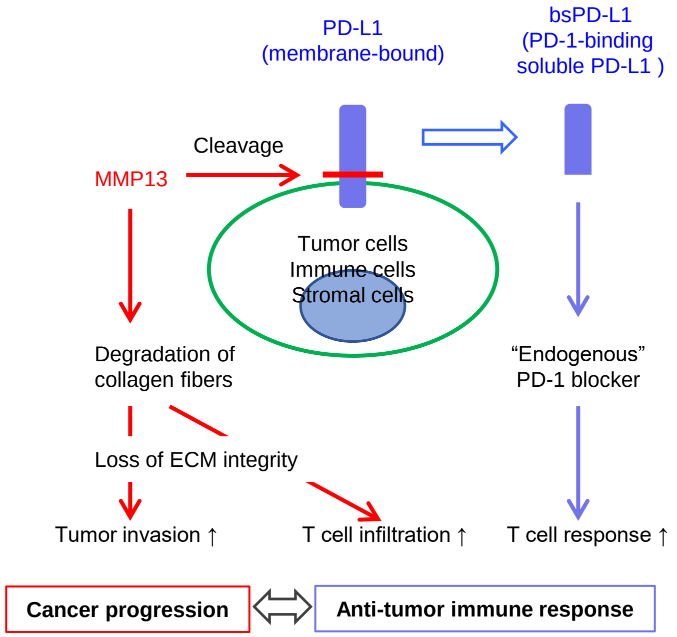


**Schematic model of the regulation of the tumor microenvironment by bsPD-L1 and MMP13 in GC patients.** MMP13 cleaves membrane-bound PD-L1 to generate bsPD-L1. bsPD-L1 binds to PD-1 and functions as an endogenous PD-1 blocker. MMP13 degrades collagen fibers in tumor tissue, resulting in loss of extracellular matrix (ECM) integrity, which causes tumor invasion as well as T cell infiltration. The balance between the anti-tumor immune response and cancer progression is important for prognosis of GC patients. High levels of MMP13 promotes tumor invasion rather than T cell response, leading to poor prognosis.
