## Supplementary Fig.6 for "Clinical significance of PD-1-binding soluble PD-L1 and MMPs in the microenvironment of gastric cancer and non-small cell lung cancer treated with PD-1/PD-L1 blockade"

### Supplementary Figure 6

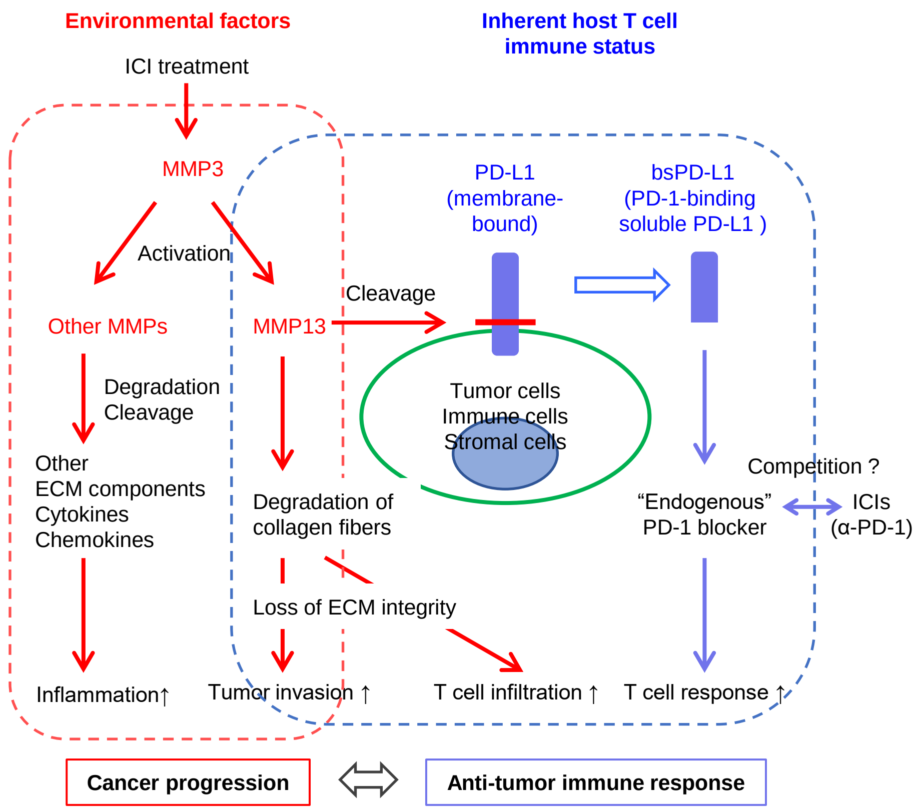


**Schematic model of the regulation of the tumor microenvironment by bsPD-L1 and MMPs in NSCLC patients with ICI treatment.** Baseline levels of bsPD-L1 or MMP13 indicate host inherent T cell immune status prior to ICI treatment. MMP13 cleaves membrane-bound PD-L1, resulting in bsPD-L1 production. bsPD-L1 may compete with ICIs such as anti-PD-1 to bind to PD-1. MMP13 and its activator MMP3 levels change during ICI treatment. MMP3 activates a wide range of MMPs and degrades a broad range of extracellular matrix (ECM) and non-matrix substrates, including cytokines and chemokines. The increase in MMP3 and MMP13 induces inflammation and accelerates cancer progression rather than T cell response. The host inherent T cell immune status and the change of MMPs (sensitivity to ICIs) vary among patients.
