## Supplementary Table1 for "Clinical significance of PD-1-binding soluble PD-L1 and MMPs in the microenvironment of gastric cancer and non-small cell lung cancer treated with PD-1/PD-L1 blockade"

**Supplementary Table 1** Characteristics of GC patients

| Variables | | All patients (n=117) | bsPD-L1-negative (n=100) | bsPD-L1-positive (n=17) | p value |
|---|---|---|---|---|---|
| Age | Median (range) | 75 (37 - 93) | 75 (37-93) | 77 (58-88) | 0.963 |
| Gender | | | | | 0.234 |
| | Male | 83 (70.9%) | 73 (73.0%) | 10 (58.8%) | |
| | Female | 34 (29.1%) | 27 (27.0%) | 7 (41.2%) | |
| Smoking | | | | | 0.831 |
| | Never | 44 (37.6%) | 38 (38.0%) | 6 (35.3%) | |
| | Current or Former | 73 (62.4%) | 62 (62.0%) | 11 (64.7%) | |
| Histology | | | | | 0.656 |
| | Differentiated | 63 (53.9%) | 53 (53.0%) | 10 (58.8%) | |
| | Undifferentiated | 54 (46.1%) | 47 (47.0%) | 7 (41.2%) | |
| T | | | | | 0.484 |
| | -1 | 36 (30.8%) | 32 (32.0%) | 4 (23.5%) | |
| | 2- | 81 (69.2%) | 68 (68.0%) | 13 (76.5%) | |
| N | | | | | 0.262 |
| | 0 | 61 (52.1%) | 50 (50.0%) | 11 (64.7%) | |
| | 1- | 56 (47.9%) | 50 (50.0%) | 6 (35.3%) | |
| M | | | | | 0.520 |
| | 0 | 105 (89.7%) | 89 (89.0%) | 16 (94.1%) | |
| | + | 12 (10.3%) | 11 (11.0%) | 1 (5.9%) | |
| Disease stage | | | | | 0.713 |
| | I - II | 71 (60.7%) | 60 (60.0%) | 11 (64.7%) | |
| | III - IV | 46 (39.3%) | 40 (40.0%) | 6 (35.3%) | |
