## Supplementary Table2 for "Clinical significance of PD-1-binding soluble PD-L1 and MMPs in the microenvironment of gastric cancer and non-small cell lung cancer treated with PD-1/PD-L1 blockade"

**Supplementary Table 2** Univariate and multivariate analysis for DFS and OS in GC patients

| | Univariate analysis for DFS | | Multivariate analysis for DFS | |
|---|---|---|---|---|
| **Variables** | **HR (95% CI)** | **p value** | **HR (95% CI)** | **p value** |
| Age ≥ 65 (vs. < 65) | 1.58 (0.67 - 4.62) | 0.320 | | |
| Male (vs. Female) | 0.75 (0.38 - 1.54) | 0.418 | | |
| bsPD-L1$^{+}$MMP13$^{high}$ (yes vs.no) | 4.19 (1.43 - 9.87) | 0.0123 | 4.05 (1.37 - 9.69) | 0.015 |
| T ≥ 2 (vs. <2) | 4.22 (1.68 - 14.16) | 0.0011 | 1.25 (0.38 - 4.92) | 0.721 |
| Lymph node metastasis (yes vs. no) | 4.32 (2.12 - 9.71) | <0.0001 | 3.46 (1.49 - 9.24) | 0.003 |
| Distant metastasis (yes vs. no) | 12.59 (5.68 - 26.20) | <0.0001 | 11.51 (4.77 - 26.87) | <0.0001 |

| | Univariate analysis for OS | | Multivariate analysis for OS | |
|---|---|---|---|---|
| **Variables** | **HR (95% CI)** | **p value** | **HR (95% CI)** | **p value** |
| Age ≥ 65 (vs. < 65) | 1.34 (0.56 - 3.99) | 0.534 | | |
| Male (vs. Female) | 0.83 (0.39 - 1.91) | 0.647 | | |
| bsPD-L1$^{+}$MMP13$^{high}$ (yes vs.no) | 4.71 (1.58 - 11.43) | 0.0082 | 2.96 (0.96 - 7.49) | 0.058 |
| T ≥ 2 (vs. <2) | 4.27 (1.51 - 17.89) | 0.0041 | 2.15 (0.58 - 10.15) | 0.257 |
| Lymph node metastasis (yes vs. no) | 3.02 (1.42 - 6.95) | 0.0036 | 1.72 (0.73 - 4.61) | 0.221 |
| Distant metastasis (yes vs. no) | 8.82 (3.53 - 20.28) | <0.0001 | 5.60 (2.11 - 13.79) | 0.001 |
