## Supplementary Table3 for "Clinical significance of PD-1-binding soluble PD-L1 and MMPs in the microenvironment of gastric cancer and non-small cell lung cancer treated with PD-1/PD-L1 blockade"

**Supplementary Table 3** Characteristics of NSCLC patients

| Variable | Variable | All patients (N=72) | bsPD-L1-negative (N=56) | bsPD-L1-positive (N=16) | p value |
|---|---|---|---|---|---|
| Age | Median (range) | 67 (28-88) | 68.5 (28-82) | 68.5 (43-88) | 0.3312 |
| Gender | | | | | 0.3247 |
| | Male | 56(77.8%) | 45(80.4%) | 11(68.8%) | |
| | Female | 16(22.2%) | 11(19.6%) | 5(22.2%) | |
| Smoking | | | | | 0.0441 |
| | Never | 11(15.3%) | 6(10.7%) | 5(31.2%) | |
| | Current or Former | 61(84.7%) | 50(89.3%) | 11(68.8%) | |
| EGFR Mutation | | | | | 0.4876 |
| | Wild | 60(88.2%) | 46(86.8%) | 14(93.3%) | |
| | Mutant | 8(11.8%) | 7(13.2%) | 1(6.7%) | |
| T | | | | | 0.5127 |
| | -1c | 18(25.0%) | 13(23.2%) | 5(31.3%) | |
| | 2a- | 54(75.0%) | 43(76.8%) | 11(68.7%) | |
| N | | | | | 0.2551 |
| | 0 | 11(15.3%) | 10(19.6%) | 1(6.3%) | |
| | 1 - | 61(84.7%) | 46(80.3%) | 15(93.7%) | |
| M | | | | | 0.9461 |
| | 0 | 49(68.1%) | 38(67.9%) | 11(68.7%) | |
| | + | 23(31.9%) | 18(32.1%) | 5(31.3%) | |
| Disease stage | | | | | 0.3164 |
| | I - II | 10 (13.9%) | 9 (16.1%) | 1 (6.3%) | |
| | III - IV | 62(86.1%) | 47 (83.9%) | 15 (93.8%) | |
| Histology | | | | | 0.7473 |
| | Squamous cell carcinoma | 32 (44.4%) | 24 (42.9%) | 8 (50.0%) | |
| | Adenocarcinoma | 32 (44.4%) | 25 (44.6%) | 7 (43.8%) | |
| | Non-small | 8 (11.2%) | 7 (12.5%) | 1 (6.2%) | |
| PD-L1 TPS status | | | | | 0.1724 |
| | <50 | 31 (52.5%) | 22 (47.8%) | 9 (69.2%) | |
| | 50≤ | 28 (47.5%) | 24 (52.2%) | 4 (30.8%) | |
