## Supplementary Table4 for "Clinical significance of PD-1-binding soluble PD-L1 and MMPs in the microenvironment of gastric cancer and non-small cell lung cancer treated with PD-1/PD-L1 blockade"

**Supplementary Table 4** Logistic regression for bsPD-L1 expression

| Variable | | AOR | 95% CI | p value |
|---|---|---|---|---|
| Smoking | | | | |
| | Never | 12.55 | 0.81 - 194.24 | 0.0703 |
| | Current or Former | 1 | | |
| TPS | | | | |
| | ≥ 50 | 0.14 | 0.01 - 2.04 | 0.1512 |
| | < 50 | 1 | | |
| Neu (/μl) | | | | |
| | ≥ 3000 | 0.15 | 0.01 - 1.82 | 0.1349 |
| | < 3000 | 1 | | |
| CRP (mg/L) | | | | |
| | ≥ 0.5 | 1.71 | 0.11 - 25.88 | 0.6983 |
| | < 0.5 | 1 | | |
| MMP13 (pg/ml) | | | | |
| | ≥ 985 | 246.35 | 10.72 - 5562.37 | 0.0006 |
| | < 985 | 1 | | |
