## Supplementary Table5 for "Clinical significance of PD-1-binding soluble PD-L1 and MMPs in the microenvironment of gastric cancer and non-small cell lung cancer treated with PD-1/PD-L1 blockade"

### Supplementary Table 5

Univariate and multivariate analysis for PFS and OS in NSCLC patients treated with PD-1/PD-L1 blockade

| | Univariate analysis for PFS | | Multivariate analysis for PFS | |
|---|---|---|---|---|
| Variables | HR (95% CI) | p value | HR (95% CI) | p value |
| Age ≥ 65 (vs. < 65) | 1.07 (0.64 - 1.83) | 0.793 | | |
| Male (vs. Female) | 0.96 (0.54 - 1.86) | 0.910 | | |
| Lymph node metastasis (yes vs. no) | 1.15 (0.59 - 2.50) | 0.704 | | |
| Distant metastasis (yes vs. no) | 0.72 (0.39 - 1.26) | 0.251 | | |
| CRP ≥ 10 (vs. < 10) | 1.52 (0.63 - 3.13) | 0.329 | | |
| NLR ≥ 3 (vs. < 3) | 0.95 (0.56 - 1.60) | 0.846 | | |
| TPS < 50 (vs. ≥ 50) | 2.35 (1.30 - 4.32) | 0.0045 | 2.31 (1.28 - 4.27) | 0.0056 |
| $MMP13^{high}$(MMP3 and MMP13) $^{increased}$ (yes vs.no) | 3.18 (1.07 - 7.72) | 0.0392 | 3.82 (1.25 - 9.73) | 0.0219 |

| | Univariate analysis for OS | | Multivariate analysis for OS | |
|---|---|---|---|---|
| Variables | HR (95% CI) | p value | HR (95% CI) | p value |
| Age ≥ 65 (vs. < 65) | 2.07 (1.05 - 4.28) | 0.0348 | 2.22 (1.12 - 4.63) | 0.0218 |
| Male (vs. Female) | 1.09 (0.52 - 2.57) | 0.821 | | |
| Lymph node metastasis (yes vs. no) | 0.72 (0.33 - 1.79) | 0.449 | | |
| Distant metastasis (yes vs. no) | 0.56 (0.25 - 1.14) | 0.112 | | |
| CRP ≥ 10 (vs. < 10) | 2.67 (1.00 - 6.02) | 0.0500 | 3.46 (1.27 - 8.04) | 0.0179 |
| NLR ≥ 3 (vs. < 3) | 1.25 (0.65 - 2.42) | 0.497 | | |
| TPS < 50 (vs. ≥ 50) | 2.06 (0.99 - 4.45) | 0.0538 | | |
| $MMP13^{high}$(MMP3 and MMP13) $^{increased}$ (yes vs.no) | 3.52 (1.02 - 9.39) | 0.0470 | 3.95 (1.13 - 10.67) | 0.0332 |
